# A model to estimate bed demand for COVID-19 related hospitalization

**DOI:** 10.1101/2020.03.24.20042762

**Authors:** Teng Zhang, Kelly McFarlane, Jacqueline Vallon, Linying Yang, Jin Xie, Jose Blanchet, Peter Glynn, Kristan Staudenmayer, Kevin Schulman, David Scheinker

## Abstract

As of March 23, 2020 there have been over 354,000 confirmed cases of coronavirus disease 2019 (COVID-19) in over 180 countries, the World Health Organization characterized COVID-19 as a pandemic, and the United States (US) announced a national state of emergency.1, 2, 3 In parts of China and Italy the demand for intensive care (IC) beds was higher than the number of available beds.4, 5 We sought to build an accessible interactive model that could facilitate hospital capacity planning in the presence of significant uncertainty about the proportion of the population that is COVID-19+ and the rate at which COVID-19 is spreading in the population. Our approach was to design a tool with parameters that hospital leaders could adjust to reflect their local data and easily modify to conduct sensitivity analyses.

We developed a model to facilitate hospital planning with estimates of the number of Intensive Care (IC) beds, Acute Care (AC) beds, and ventilators necessary to accommodate patients who require hospitalization for COVID-19 and how these compare to the available resources. Inputs to the model include estimates of the characteristics of the patient population and hospital capacity. We deployed this model as an interactive online tool.^6^ The model is implemented in R 3.5, RStudio, RShiny 1.4.0 and Python 3.7. The parameters used may be modified as data become available, for use at other institutions, and to generate sensitivity analyses.

We illustrate the use of the model by estimating the demand generated by COVID-19+ arrivals for a hypothetical acute care medical center. The model calculated that the number of patients requiring an IC bed would equal the number of IC beds on Day 23, the number of patients requiring a ventilator would equal the number of ventilators available on Day 27, and the number of patients requiring an AC bed and coverage by the Medicine Service would equal the capacity of the Medicine service on Day 21.

In response to the COVID-19 epidemic, hospitals must understand their current and future capacity to care for patients with severe illness. While there is significant uncertainty around the parameters used to develop this model, the analysis is based on transparent logic and starts from observed data to provide a robust basis of projections for hospital managers. The model demonstrates the need and provides an approach to address critical questions about staffing patterns for IC and AC, and equipment capacity such as ventilators.

---

As of March 23, 2020 there have been over 354,000 confirmed cases of coronavirus disease 2019 (COVID-19) in over 180 countries, the World Health Organization characterized COVID-19 as a pandemic, and the United States (US) announced a national state of emergency.^1, 2, 3^ In parts of China and Italy the demand for intensive care (IC) beds was higher than the number of available beds.^4, 5^ The limited availability of testing results in the US make it challenging to estimate the demand for hospital beds that will be generated by COVID-19 patients. We sought to build an accessible interactive model that could facilitate hospital capacity planning in the presence of significant uncertainty about the proportion of the population that is COVID-19+ and the rate at which COVID-19 is spreading in the population. Our approach was to design a tool with parameters that hospital leaders could adjust to reflect their local data and easily modify to conduct sensitivity analyses.

We developed a model to facilitate hospital planning with estimates of the number of Intensive Care (IC) beds, Acute Care (AC) beds, and ventilators necessary to accommodate patients who require hospitalization for COVID-19 and how these compare to the available resources. We deployed this model as an interactive online tool.^6^

Inputs to the model include estimates of the characteristics of the patient population and hospital capacity. The patient population inputs are the starting population of COVID-19-patients, COVID-19+ patients, and the estimated doubling time for total COVID-19 admissions (i.e. how many days it will take for the number of total admissions at the institution to double). For the patient cohorts, we estimated what percentage of the COVID-19+ patient population would follow 5 potential trajectories through the hospital, assuming various combinations of patient flow between IC and AC. Until more reliable data are available, estimates for input values are generated through review of existing data and discussion with experts.^7, 8^ The model will be updated as data become available.

The model produces a daily time series. The first day of the simulation (Day 0) is fixed. For each subsequent day the model uses the projected number of COVID-19 patients, partitions the patients into cohorts, and updates the number of COVID-19 patients requiring IC and AC beds as follows:

1. <u>COVID-19 Admissions:</u> We project the total COVID-19 admissions with an exponential growth model using data based on the first 14 days of patient admissions (The number of total admitted patients up to day n is the product of the number of patients admitted up to day 0 and 2 to the power of n divided by the doubling time).
2. <u>Patient Cohorting:</u> The patients are partitioned into 5 care cohorts: Cohorts 1 and 5 are patients who spend time only on, respectively, a General Medicine AC floor and an IC unit. The remaining 3 cohorts are patients who spend time in an IC unit, only before, only after, or both before and after spending time on an AC unit (Table 1).
3. <u>Cohort Length of Stay:</u> Each patient in each cohort spends the number of days specified by the parameter inputs in the IC and AC units. The total census in each of the IC and AC units for each day is calculated as the sum of the patients that arrived in that unit minus the sum of the patients that have been discharged from that unit.
4. <u>Projected IC Bed Requirements:</u> The number of IC beds required each day is the sum of the number of COVID-19+ and COVID-19-IC patients.
5. <u>Projected COVID-19 Medical Team Requirements:</u> The number of patients to be cared for by the Medical Service each day is the sum of the number of COVID-19+ AC patients and COVID-19-patients being cared for by the Medicine Service.
6. <u>Projected AC Bed Requirements:</u> The number of AC beds required each day is the sum of the number of COVID-19+ and COVID-19-AC patients.
7. <u>Ventilator Requirements:</u> The number of ventilators required is estimated as the sum of 50% of non-COVID-19 IC patients and 100% of COVID-19+ IC patients.

The model is implemented in R 3.5, RStudio, RShiny 1.4.0 and Python 3.7. The parameters used may be modified as data become available, for use at other institutions, and to generate sensitivity analyses.

**Table 1:** Patient Cohorts and Length of Stay Estimates.

| COVID Patient Cohorts |  |  | LOS Estimates in Model |  |  |  |
| --- | --- | --- | --- | --- | --- | --- |
| Index | Path | Fraction of Patients (%) | LOS Floor | LOS ICU | LOS Floor | Total LOS |
| 1 | Floor | 70.4 | 5 |  |  | 5 |
| 2 | Floor -> ICU -> Floor | 13.0 | 4 | 9 | 4 | 17 |
| 3 | Floor -> ICU | 1.8 | 6 | 9 |  | 15 |
| 4 | ICU -> Floor | 13.0 |  | 9 | 4 | 13 |
| 5 | ICU | 1.8 |  | 11 |  | 11 |

We illustrate the use of the model by estimating the demand generated by COVID-19+ arrivals for a hypothetical acute care medical center with 100 IC beds, 220 AC beds, 75 ventilators, 80% occupancy of both IC and AC beds, 1 COVID-19+ IC patient, 2 COVID-19+ AC patients, and a total patient doubling time of 6 days. For COVID-19+ patients, 70% were assumed to stay only in AC for an average LOS of 5 days and 30% were assumed to spend at least one day in IC with an average IC LOS of 8 days and average AC LOS of 9 days (Table 1).

## Model projections

The model calculated that the number of patients requiring an IC bed would equal the number of IC beds on Day 23 (Figure 1a), the number of patients requiring a ventilator would equal the number of ventilators available on Day 27 (Figure 1a), and the number of patients requiring an AC bed and coverage by the Medicine Service would equal the capacity of the Medicine service on Day 21 (Figure 1b).

**Figure 1:**
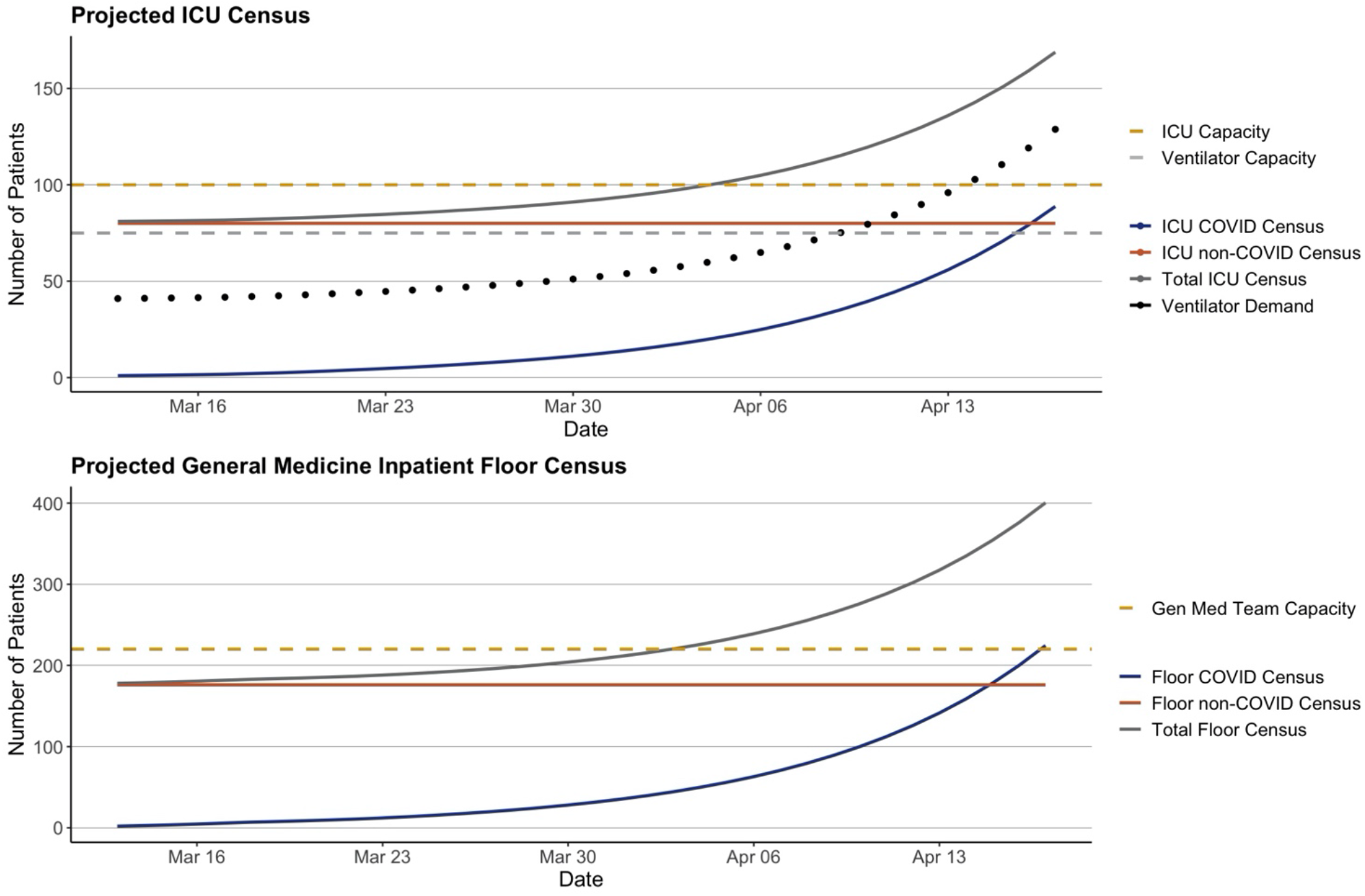
Projected Total ICU and General Medicine Team Acute Care Census.

## Sensitivity analyses

When the doubling time of new admissions is decreased to 3 days (50%), the number of days until IC and AC hit capacity decrease are, respectively, 11 and 9. When the doubling time is increased to 12 days (200%), the number of days until IC and AC hit capacity are, respectively, 51 and 51.

## Discussion

In response to the COVID-19 epidemic, hospitals must understand their current and future capacity to care for patients with severe illness. While there is significant uncertainty around the parameters used to develop this model, the analysis is based on transparent logic and starts from observed data to provide a robust basis of projections for hospital managers. The model demonstrates the need to address critical questions about staffing patterns for IC and AC, and equipment capacity such as ventilators.

An insight revealed by the model is that under some plausible scenarios, AC may reach capacity before IC and become a bottleneck preventing discharges from IC. In addition to increasing capacity, managers must develop strategies to reduce AC occupancy such as accelerating efforts to discharge patients to convalescent or step-down care such as a hotel or nursing care facility.

The main limitation of this model is the fact that most of the inputs are based on estimates. The epidemiology of COVID-19 is critically important, and ongoing research will update the model. The model is very sensitive to specific aspects of the epidemiology, especially doubling time. The model environment can be easily updated with new parameter data to generate a more precise projection.

## Data Availability

The model referenced in the paper is available online at the link included below. All model data are available on the Tab titled "Tabular Representation".

https://surf.stanford.edu/covid-19-tools/covid-19-hospital-projections/

## Author Statement

The authors have no sources of funding nor conflicts or other disclosures. Teng Zhang, Kelly McFarlane, and Jacqueline Vallon have reviewed all of the data and analyses and take responsibility for their accuracy. Teng Zhang, Kelly McFarlane, Jacqueline Vallon, Linying Yang, and Jin Xie built the model. Jose Blanchet, Peter Glynn, Kristan Staudenmayer, Kevin Schulman, and David Scheinker led the conceptualization and design of the model. Teng Zhang, Kelly McFarlane, Kevin Schulman, and David Scheinker led drafting the correspondence. All authors contributed to critical reading and revision of the correspondence.

This is a mathematical model with no human subjects data.

